# Exploring the potential risk factors of myopia: A phenome-wide Mendelian randomization association study

**DOI:** 10.1101/2024.06.24.24309385

**Authors:** Di Hu, Junhong Jiang, Qi Zhang, Zenan Lin, the μ-Biomedical Data Investigation Group (Mu-BioDig)

## Abstract

**Introduction:** Myopia is a leading cause of visual impairment worldwide, whose pathogenesis remains poorly understood. Identification of its risk factors would benefit the management and treatment of myopia.

**Methods:** We comprehensively performed phenome-wide Mendelian randomization analysis (MR-PheWAS) to explore causal factors and potential therapeutic targets for myopia in participants from the UK Biobank study.

**Results:** Our PheWAS revealed that 55 robust associations (1 disease, 2 employment, 3 cognitive functions, 4 sex-specific factors, 4 mental health factors, 5 lifestyle and environmental factors, 10 sociodemographic factors, 12 physical measures and 14 ocular measures/conditions) were significantly causally correlated with myopia.

**Conclusions:** The results indicate that myopia may be influenced by several factors, including serum metabolic traits, fatty acid intake, fat-related indices, mental health, as well as some previously acknowledged risk factors. Future clinical trials are needed to verify our results.

## 1. Introduction

Myopia, characterized by progressive and irreversible axial length elongation ^1^, is becoming a prominent eye disease worldwide. It is estimated that 50% of the global population suffers from myopia, with a continuously increasing number in 2050, imposing a substantial economic burden of as much as $1.7 trillion.^2^ Besides blurred vision caused by optical myopic defocusing, the continuous growth of the axial leads to high myopia, which is believed to be associated with a high risk of irreversible visual impairments, including glaucoma, macular degeneration, and retinal detachment.^3^ Myopia is becoming a significant global health concern, but effective methods of myopia control remain limited because of its complicated pathogenesis.

The pathogenesis of myopia is highly complex and multifactorial, involving an interaction between genetic and environmental factors.^4,5^ Variations in almost 100 genes and mutations in more than 20 chromosomal locis have been reported to be associated with myopia.^6^ In addition to genetic factors, environmental factors are linked to the risk of myopia, including near work, screens of computers and handheld devices, educational stress, family income, and living environments.^7^ However, most evidence has come from observational studies, which are subject to bias and may not confirm causal effects.

Phenome-wide Mendelian randomization (MR) is now widely used to look for the comprehensive causality between exposures and disease outcomes using hypothesis-searching.^8^ The method is typically less prone to residual confounding and reverse causation than observational studies,^9^ which are used as an alternative to randomized clinical trials. In this study, we performed a phenome-wide MR study to investigate the relationship between myopia and more than six thousand exposure variables.

## 2. Methods

### 2.1 Study design and data sources

The overall study design is outlined in Figure 1. We first conducted an MR-phenome-wide association study (MR-PheWAS) to explore the potential factors for myopia in the UK Biobank study. The UK Biobank is a large prospective population-based cohort study that recruited over 500,000 participants aged between 40 and 69 years across the United Kingdom between 2006 and 2010.^10^ The myopia phenotype GWAS summary dataset of the UK Biobank was acquired from the MRC Integrative Epidemiology Unit (IEU) OpenGWAS database (https://gwas.mrcieu.ac.uk/).^11^ The basic features and the detailed quality control procedures of phenotype data are described Table S1.

**Figure 1.**
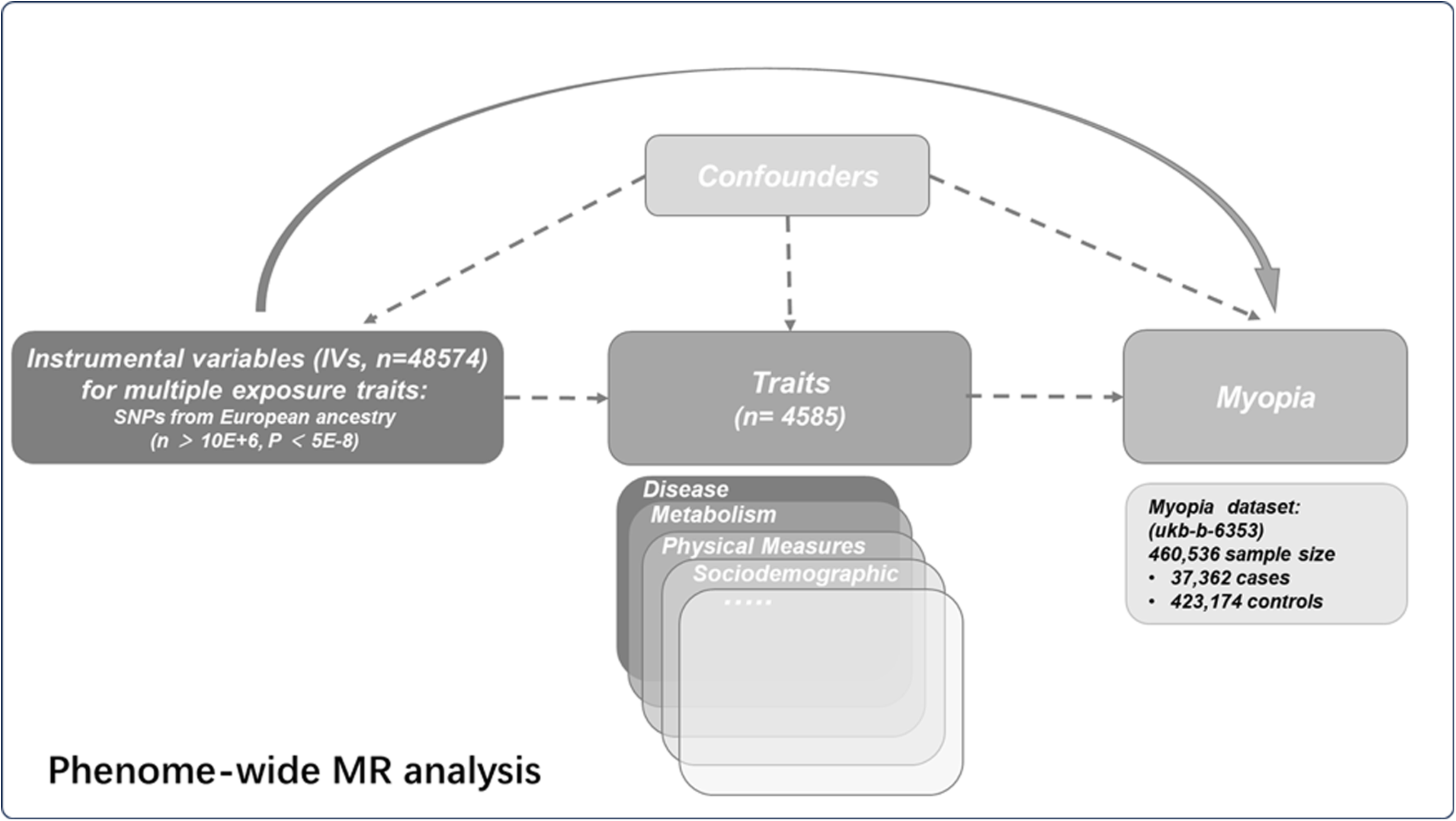
The overall study design of the current study.

### 2.2 Genetic instrument selection

We acquired all the available GWAS datasets in the IEU GWAS platform from inception to May. 06. 2023. During the selection process, the datasets analyzed only in European participants were kept. In order to control the quality of the datasets, those with a sample size less than 3000 or SNPs less than 10E+6 were also excluded for further analyses. ‘ukb-a’ associated traits were excluded since the new version ‘ukb-b’ was available.

### 2.3 Phenome-wide MR analysis

Then, the traits were classified into the following 19 categories according to their features: fMRI, proteins, metabolites, physical measures, diseases, lifestyle and environment, ocular measure/condition, treatment/medication, family factor, sociodemographics, health conditions, mental health, smoking, alcohol drinking-associated, employment, gut microbiota abundance, cognitive function, sex-specific factor and others. All selected SNPs met the following criteria: 1) P values < 5E-8; 2) linkage disequilibrium (LD) clumping was performed with the PLINK (version 1.9)^12^ to find independent SNPs (r2 < 0.001 within 10Mb), and the 1000 Genomes Project Phase 3 (EUR) was utilized as the reference panel; 4) those with lowest P values were retained when duplicated SNPs were found; 5) the proxy SNPs (r2 > 0.8) were used when the SNPs were not found in the outcome’s dataset. In the end, 103433 IVs for 6481 traits were kept for further MR investigations (See Table S2.). As some traits may be associated or even duplicated, the management of these traits is a problem. Since there was no consensus on selecting a perfect representing trait, they were all included in the analyses. Finally, a conservative Bonferroni-corrected *P* threshold was applied to verify the findings. After harmonizing the data of exposures and outcome, the MR investigations were successfully performed on 6273 valid exposure traits (See Table S3.).

### 2.4 Statistical analysis

In summary, in the MR analyses between various phenotypical traits and myopia, the Wald ratio (for single IV) and inverse-variance-weighted (IVW, for IVs ≥ 2 SNPs) methods were introduced as the primary analytical tools. When the IVW analysis was conducted, complementary analyses such as the simple mode, weighted median, weighted mode, and MR-Egger regression were introduced to assess the robustness of identified causality between the exposures and outcome. The analyses were conducted with R package TwoSampleMR (version 0.5.6)^11^, plinkbinr (version 0.0.0.9000), ieugwasr (version 0.1.5), gwasglue (version 0.0.0.9000). The statistically significant *P* threshold was set at the Bonferroni-corrected level whenever applicable.

## 3. Results

### 3.1 PheWAS Analyses Identified 852 Factors Spanning 19 Categories as Causal Influencers on Myopia

After the MR analyses, a total of 852 traits showed significant casual effects on myopia (Figure 2 and Table S4.), including 389 positive and 463 negative associations. Among these, 204 were fMRI-associated traits. The others included 145 proteins, 99 metabolites, 71 physical measures, 70 diseases, 51 lifestyle and environmental factors, 33 ocular measures/conditions, 33 treatment/medications, 23 family factors, 17 sociodemographics, 13 health conditions, 12 mental health, 12 smoking-related factors, 10 alcohol drinking-associated factors, 9 employment factors, 8 gut microbiota abundances, 5 cognitive functions, and 4 sex-specific factors (and Table S4.).

**Figure 2.**
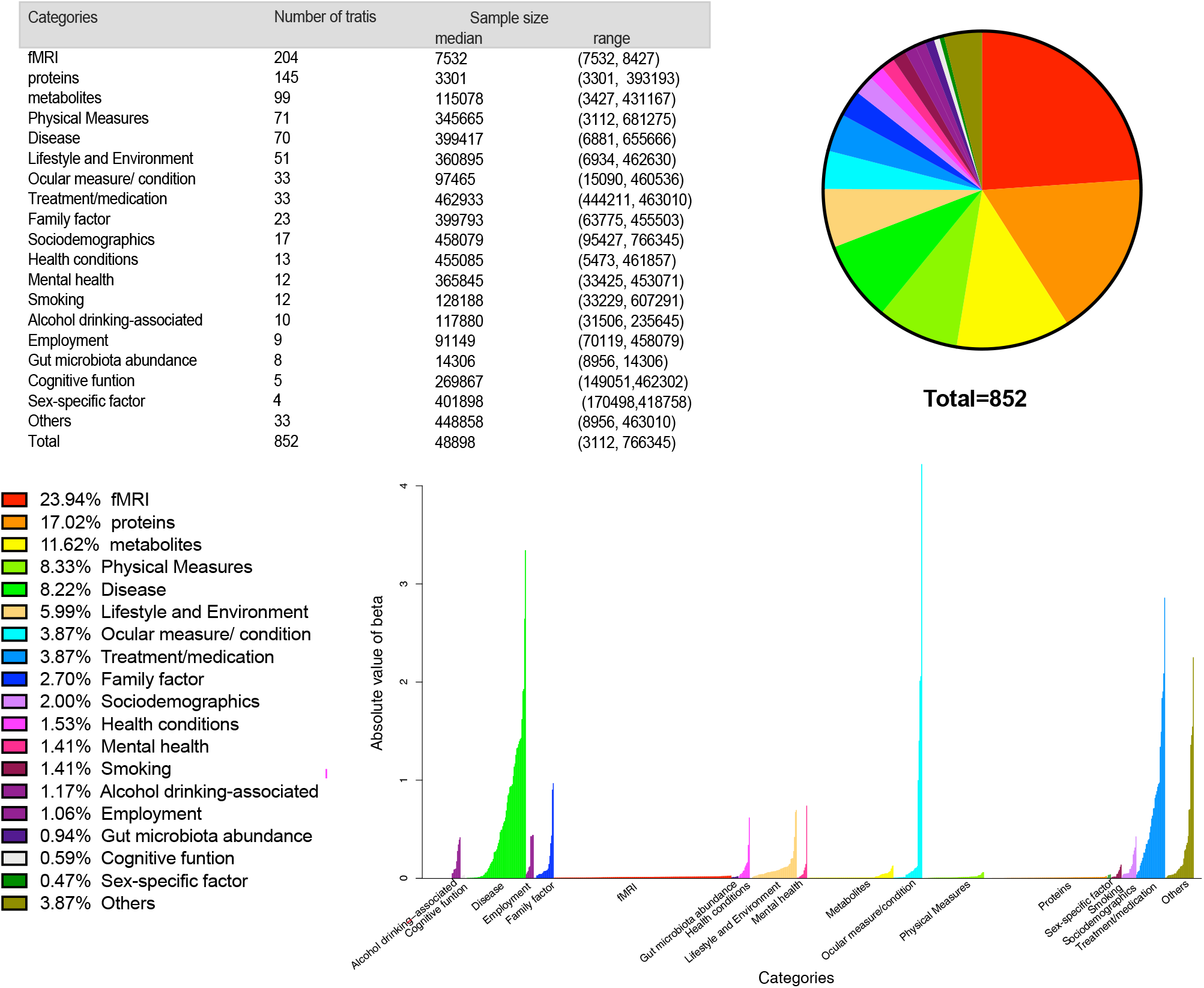
Overview of risk factors. There were 852 factors from 19 categories included.

### 3.2 Robust factors

To clarify factors which has the most significant effect on myopia, we performed phenotype screening using a *P* < 0.05/6273 as cut-off. As shown in Fig. 3 and Table S5., the most robust findings were identified for 55 traits, including 1 disease, 2 employment factors, 3 cognitive functions, 4 sex-specific factors, 4 mental health traits, 5 lifestyle and environmental factors, 10 sociodemographic factors, 12 physical measures, and 14 ocular measures/conditions.

**Figure 3.**
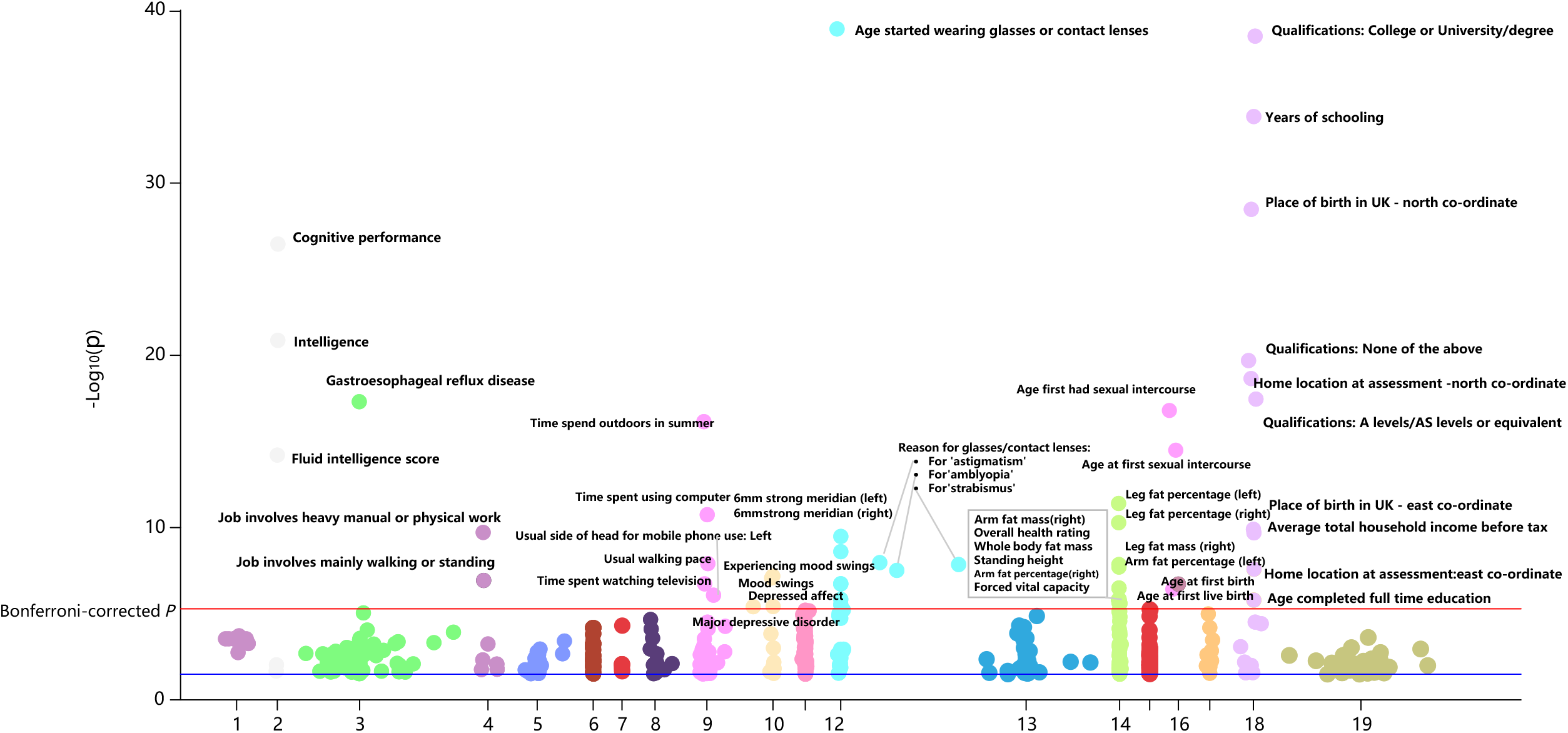
55 robust phenotypes demonstrate remarkable causal impact myopia. Those with *P* values less than the Bonferroni-corrected level were highlighted.

### 3.3 Metabolites associated with myopia risk

In the 99 serum metabolic associations identified as potential causal effects (see Table S6.), 33 were potentially risk and 66 were protective factors for myopia. These serum metabolic traits included fatty acids, lipids, lipoproteins, cholesterol, glycerides, and phospholipids measures, amino acids, ketone bodies, glycolysis-related metabolites, and circulating protein.

As shown in Figure 4. and Table S7., we further analyzed the most significant factors associated with myopia using the MR approach based on Bayesian model averaging (MR-BMA). The top 10 individual risk factors in terms of marginal inclusion probability are eicosapentaenoate (EPA), urate acid, glutaroyl carnitine, albumin, free cholesterol to total lipids ratio in very small VLDL, free cholesterol to total lipids ratio in very large VLDL, acetoacetate, 1-oleoylglycerol (1-monoolein), phospholipids to total lipids ratio in very large VLDL, and phospholipids to total lipids ratio in medium VLDL. (See Table S7.)

**Figure 4.**
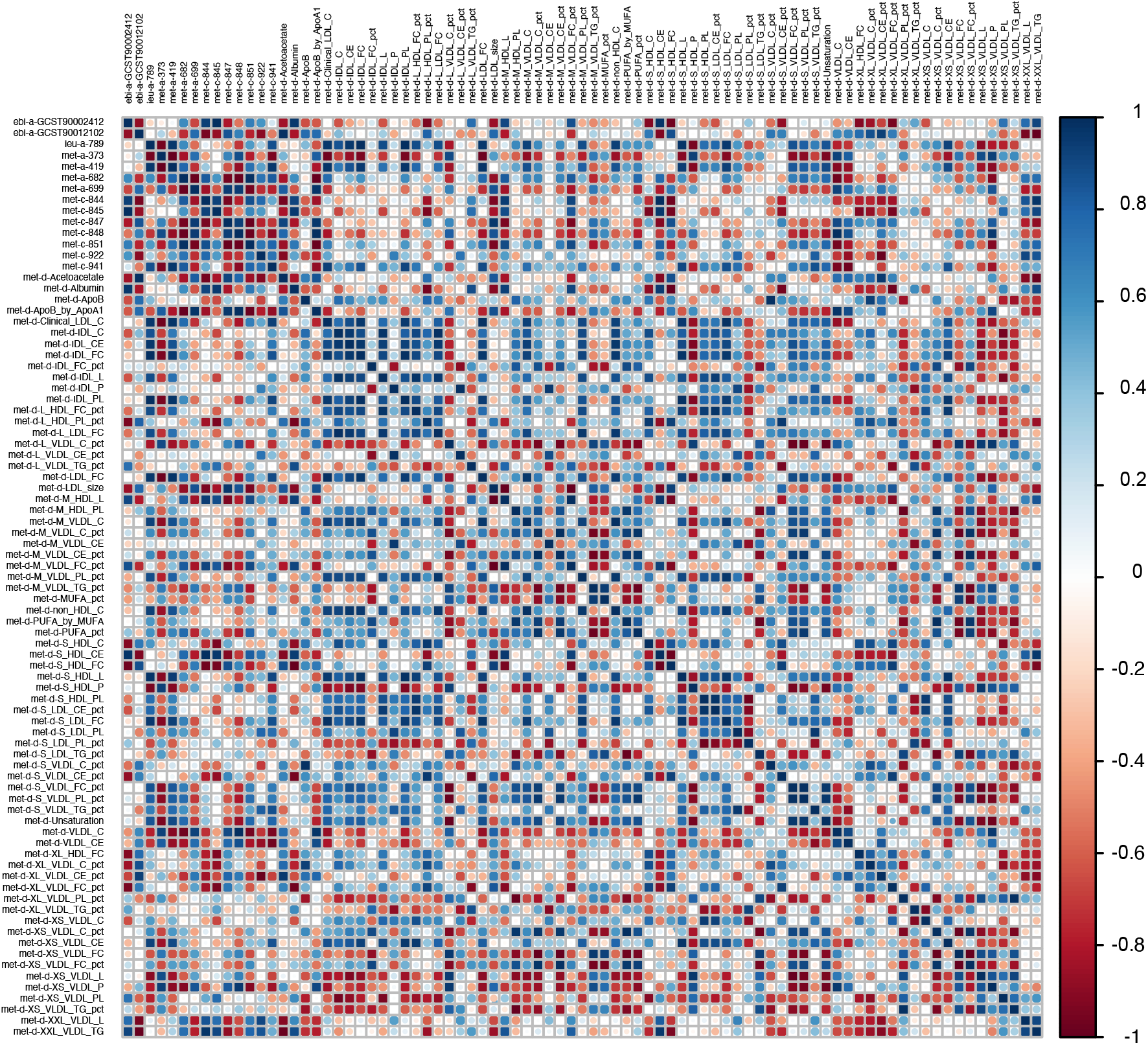
The plot illustrating the correlations between the beta values for the metabolites considered in our Mendelian randomization Bayesian model averaging (MR-BMA) analysis for myopia.

### 3.4 Diseases impacting myopia risk

Seventy disease traits were associated with genetically predicted myopia, with 28 being risk factors and 42 being protective factors (See Figure 5 and Table S8.). Disease phenotypes/groups with five highest positive causal effect on myopia included glaucoma (Beta=0.126199576, *P*=1.38E-05), macular degeneration (Beta=0.262906638, *P*=0.000136162), bilateral inguinal hernia without obstruction or gangrene (Beta=3.343691183, *P*=0.000181818), type 2 diabetes (Beta=0.003737744, *P*=0.00041539) and early age-related macular degeneration (Beta=0.003899109, *P*=0.000589795). Traits with the highest negative causal impact were gastroesophageal reflux disease (Beta=-0.021679083, *P*=7.58E-18), osteoarthritis of the hip or knee (Beta=-0.011080809, *P*=0.000528061), meniscus derangement (Beta=-0.469568121, *P*=0.001390964), other joint disorders (Beta=-0.326926084, *P*=0.00162895) and hallux valgus (Beta=-0.865267706, *P*=0.002337164).

**Figure 5.**
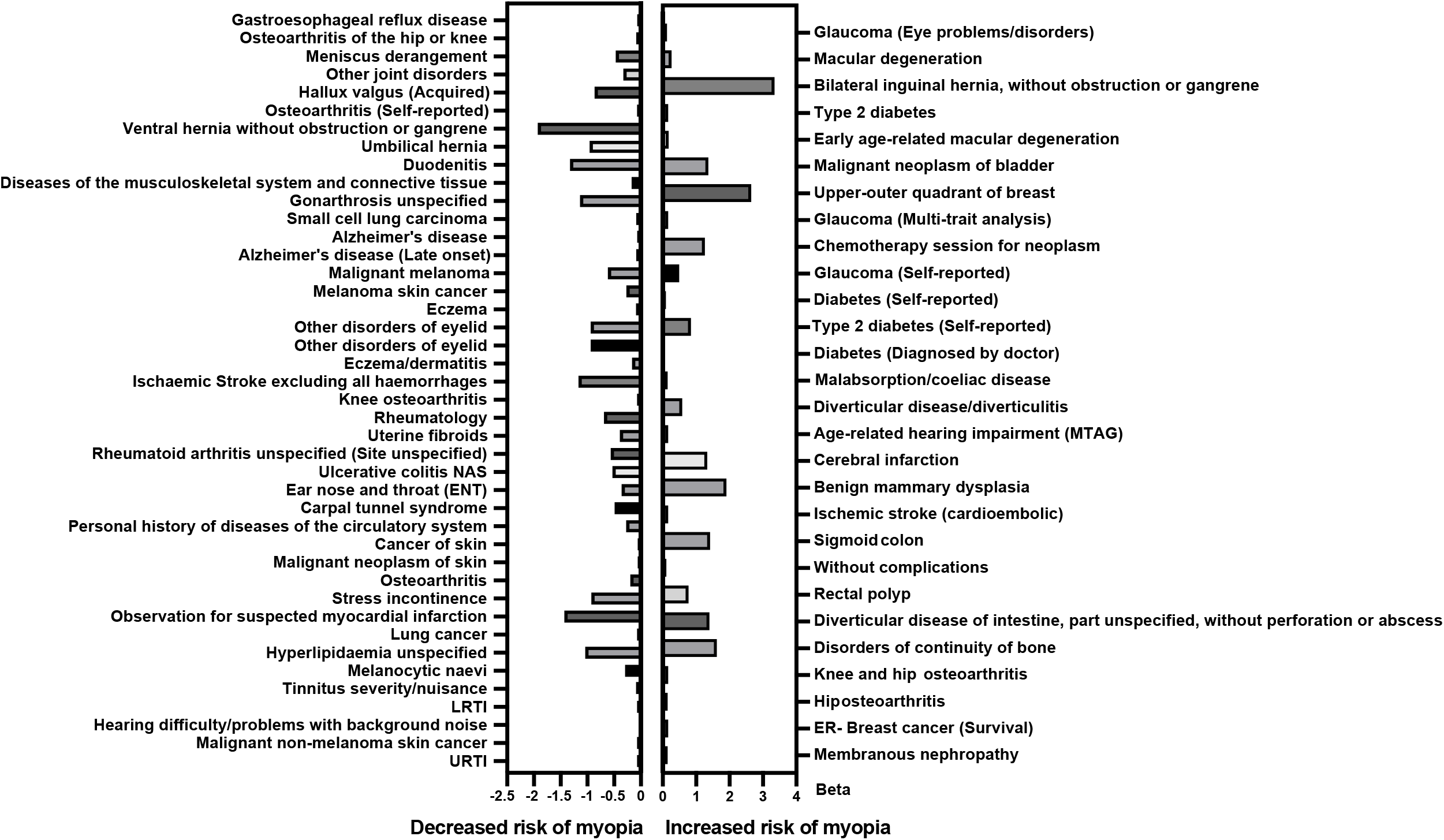
Disease phenotypes with significant causal influences on myopia risk.

### 3.5 Lifestyle and Environment

The outdoor time in summer, the time and frequency of physical activity (walking, moderate to vigorous activity) were negatively associated with the risk of myopia, and intake of red wine, beer plus cider, oily fish, vitamin B6, decaffeinated coffee, and variation in diet reduced the risk of myopia (See Figure S1. and Table S9.). Interestingly, we found that the time spent watching television had a significantly negative correlation with myopia. This is supported by the observation of a long-term follow-up RCT study that those who spent more time watching television were less likely to develop myopia than those who watched television <3 hours daily. ^13^ Positive correlations were also observed, such as time spent using a computer, the usual side of the head for mobile phone use, the usual walking pace, particulate matter and nitrogen dioxide air pollution.

### 3.6 Physical Measures

As shown in Figure S1. and Table S10., numerous physical measures showed causal effects on myopia, such as body stature traits, fat-related traits and immunophenotypes. Of the body stature traits, height, standing, and sitting height were positively correlated with myopia, while body mass index, waist circumference, hip circumference, and weight were significantly negatively correlated with myopia. Regarding the body composition parameters, the percent liver fat, trunk, and leg fat-free mass, and whole body fat-free mass were positively associated with myopia, while the leg fat percentage, body fat percentage, leg fat mass, arm fat percentage, arm fat mass, whole body fat mass, trunk fat percentage, trunk fat mass, and pancreas fat were negatively correlated with myopia (See Figure S1. and Table S10.).

In terms of immunophenotypes, anti-Merkel cell polyomavirus IgG seropositivity, IgD^-^ CD27^-^ B cell %lymphocyte, CD19 on IgD^+^ CD24^+^ B cell, FSC-A on CD8^+^ T cell, CD19 on IgD^-^ CD38dim B cell, CD19 on switched memory B cell, CD19 on B cell were positively associated with myopia (See Figure S1 and Table S10.). And IgD on IgD^+^ CD38^-^ B cell, IgD on unswitched memory B cell, HLA DR^++^ monocyte %leukocyte, HLA DR on CD14^+^ CD16^-^ monocyte, HLA DR on CD14^+^ monocyte, CD27 on memory B cell, CD45RA^-^ CD28^-^ CD8^+^ T cell %T cell, CD27 on unswitched memory B cell and Terminally Differentiated CD4^+^ T cell %T cell were positively associated with myopia. Other factors positively associated with increased myopia risk included lung function traits, such as forced vital capacity, fluid expiratory volume and peak expiratory flow, and brain intracranial volume.

### 3.7 Sociodemographics Traits

As expected, educational phenotypes including college or university degree, college completion, years of schooling, and age completed full-time education were significantly positively causally correlated with myopia (See Figure S1. and Table S11.), whereas year ended full-time education was negative. Moreover, birthplace and residential environment were strongly associated with myopia; born or resident in the east region of the UK were positive, whereas born or resident in the north area of the UK, rent in local authority local council housing association were negatively correlated with myopia. Average total household income before tax was positively correlated with myopia.

### 3.8 Other factors

Of note, other traits such as mental health, smoking-related phenotypes, alcohol phenotypes, sex-specific factors, and treatment/medications demonstrated causal impacts on myopia (See Figure S1. and Table 12-16.). Bipolar disorder showed significant positive correlations with myopia. Experiencing mood swings, mood swings, depressed affect, major depressive disorder, neuroticism, feeling fed-up, neuroticism, anxiety nerves, or generalized anxiety disorder were causally negative with myopia. Notably, age at first sexual intercourse and age at first live birth were positively associated with a higher risk of myopia.

## 4. Discussion

To our knowledge, this is the first study to comprehensively evaluate the causality between a wide array of risk factors and myopia by integrating genetic, clinical, behavioral, environmental, sociodemographic and health information data comprising 6273 traits. Overall, 852 traits showed significant causal effects on myopia, which 55 met the stringent our criteria of a Bonferroni-corrected approach. Therefore, our study has offered a novel perspective to understand the mechanism of myopia and provides important insights for the prevention and treatment of myopia. Metabolic factors play important roles in the pathogenesis of myopia. Based on Bayesian network averaging analysis, we found that eicosapentaenoate (EPA), urate acid, glutaroyl carnitine, albumin, free cholesterol to total lipids ratio in very small VLDL, free cholesterol to total lipids ratio in very large VLDL, acetoacetate, 1-oleoylglycerol (1-monoolein), phospholipids to total lipids ratio in very large VLDL, and phospholipids to total lipids ratio in medium VLDL were directly related to myopia. EPA, an ω-3 C20-polyunsaturated fatty acid with a wide range of functions in biological systems, was robustly associated with myopia. The detected effect size highlighted that genetic liability to increasing serum EPA levels is protective of myopia (beta=-0.060657022, *P* value= 0.000434288). Intriguingly, the intake of EPA was recently demonstrated to delay the development of myopia in experimental animals significantly.^14^ Uric acid (UA), a protective substance against oxidative damage in the central nervous system, was found to be at significantly lower in primary open-angle glaucoma patients versus controls.^15^ Recently, one case-control study observed that uric acid was closely correlated with pathological myopia.^16^ Additionally, enhanced levels of albumin were observed in the aqueous humor of patients with high myopia.^17^ The direct evidence on the associations between the other 7 factors and myopia is limited. Thus, more relevant studies are warranted to verify our findings.

Genetic correlation analyses indicate that myopia shares genetic liability with several diseases, such as eye disease, metabolic disease, neurological disease, osteoarticular disorders, digestive system diseases, and immune-related diseases. The strongest associations in eye disease were glaucoma and macular degeneration.^18,19^ A recent MR study indicates that there is a solid bidirectional genetic causality between myopia and primary open-angle glaucoma and IOP is the major mediator.^20^ The result is consistent with our study. Our study first demonstrates the causality between myopia and AMD. Previous study on the association between myopia and type 2 diabetes mostly came from cohort studies and cross-sectional studies. The changes of hyperglycemia in lens are recognized to cause myopia. ^21^ Similarly, our study suggested diabetes was to be a risk factor of myopia. Furthermore, diseases such as hallux valgus, osteoarthritis, hernia, diverticular disease/diverticulitis, joint disorders, ventral hernia, Stickler syndrome, Donnai-Barrow syndrome, and diverticular disease/diverticulitis were also found to have causal impacts on myopia.^22^ To our knowledge, they were rarely reported to be associated with myopia. These factors may be worthwhile to investigate in further myopia studies.

Lifestyle factors were widely acknowledged to play a vital role in one’s susceptibility to myopia. In our study, life habits, including sports, dietary habits, time spent outdoors in summer, and use of computers, mobile phones, and television, are observed to be related to myopia risk. It is not unexpected that the increased use of computers and mobile phones was positively associated with the risk of myopia, which is in line with previous studies.^23^ Interestingly, a negative correlation between the time spent watching television and myopia was observed in this study, which is in accordance with the observation of a 23-year follow-up RCT study that those who spent more time watching television were less likely to have myopia than those who watched television <3 hour daily.^13^ Our study supports the findings of previous research that outdoor time in summer, the time and frequency of physical activity (walking, moderate to vigorous activity) were negatively associated with the risk of myopia. Increasing outdoor time was shown to be effective in reducing the risk of myopia onset and myopic progress in previous report.^24^ Debate exists over the relationship between physical activity and the risk of myopia.^25^ No significant association was observed between physical activity and myopia in the study of a Danish child cohort.^26^ In contrary, our reported association of physical activity with myopia risk, supporting findings from an observational study with a 1443 child cohort, that lower physical activity was associated with higher odds of myopia,^26^ highlighting the importance of increasing physical activity as a way for myopia prevention.

The association between dietary habits and myopia has been previously reported.^27^ In this study, we discovered that the high intake of oily fish may decrease the risk of myopia. Oily fish is the primary dietary source of omega (ω)-3 long-chain polyunsaturated fatty acids; supplementation with ω-3 long-chain polyunsaturated fatty acids has been indicated to inhibit myopia progression in animal and human studies, which was in accordance with our study.^14^ Other nutrient-related traits, including vitamin B6, decaffeinated coffee, salt, alcohol, red wine, beer plus cider, and variation in diet were also identified to reduce the myopia risk. Although the protective effects of vitamin B6 have been demonstrated in age-related macular degeneration (AMD) and diabetic retinopathy (DR),^28,29^ its role in myopia has not yet been explored. Resveratrol, as the major substance in red wine, can improve microcirculation and is considered to prevent glaucoma, AMD, and DR. However, the association between red wine and myopia has not yet been reported.

Meanwhile, our study found that several sociodemographic factors are associated with the risk of myopia, which agreed with the common views of previous investigations. Studies over the past hundred years have established the influence of education on myopia risk. We observed that education (including traits such as years of schooling, age completed full-time education, and college/university degree) is a causal risk factor for myopia. Consistently, previous literature has indicated that higher education was associated with more severe myopia.^30^ Education is universally identified as the most important environmental risk factor for myopia, and the rate was about 4 times higher in college graduates than in persons with primary schooling.^5^ The age-standardized prevalence of myopia in persons with primary, secondary, and higher education was respectively 25.4%,29.1%, and 36.6%.^30^ Longer near-work time is required for a higher level of education, which is hypothesized to be responsible for the progression of myopia.^31^ Studies have shown that the effect of the myopia gene may be influenced by education. Aside from conventional myopia-related genes, like *MMP1* and *MMP10*,^32^, people with genetic variants near education-associated genes (e.g. *GJD2, RBFOX1, LAMA2, KCNQ5*, and *LRRC4C)* were confirmed susceptible to myopia.^33^ Whether the education-myopia interaction was mediated by these genes is an interesting question that requires further investigation.

The relationship between socioeconomic status and myopia has been the subject of debate. Financially constrained individuals have been shown to be more prone to suffer from myopia in the Generation R study.^34^ Further, a recent study in Shanghai showed that higher family income was negatively correlated with the risk of myopia in children.^35^ However, other studies demonstrated that the population with higher socioeconomic status has a greater prevalence of myopia.^36^ Our results show that household income positively contribute to the myopia risk. We speculated that increased parental enthusiasm for education in high-income households may be responsible for the high myopia incidence, resulting in their kids being forced to spend more time near homework.

Our findings support that height, weight, and BMI play important roles in myopia pathogenesis. Prior investigation has confirmed that taller height was associated with higher myopia.^37^ Myopic males were found to be 1.9 cm taller than nonmyopic males in a Finnish study.^38^ Another Danish study showed that myopes were 0.8 cm taller than emmetropes.^39^ However, a study with 106,926 adults in Israel found no relationship between height and myopia.^40^ The inconsistent research findings may be a result of race/ethnic differences. In South Korea, a study of 33,355 Koreans identified that weight was positively associated with myopia in children. However, there is no correlation between weight, waist circumference, BMI, and myopia prevalence in adults.^37^ Our results show a positive association between height and myopia, while a negative association between weight, BMI, and myopia.

This study has several strengths. Firstly, we used a PheWAS approach to comprehensively evaluate the causality between multiple phenotypes and myopia, which is less constrained by prior assumptions based on an incomplete understanding of disease mechanisms and less susceptible to reverse causality than traditional observational studies. Furthermore, the analysis presented here is based on the combined large-scale GWAS from the well-represented cohorts. Due to the exploratory nature of this research project, some potential limitations of this study deserve consideration. A major limitation is that MR cannot fully recapitulate a clinical trial. Second, the true risk factors on myopia are likely to vary by life stage and be more complex than indicated by our study. Third, the analyzed dataset with participants majorly being European may only partially represent the general population. Therefore, it is warranted that the current findings should be validated in additional larger samples. Fourth, due to the updated access policy of IEU GWAS platform, a large scale in-parallel PheWAS analysis was not possible anymore, thus, the exposure traits analyzed were only from inception to May. 06. 2023. Fifth, myopia was analyzed only as outcome in the PheWAS, thus, potential bidirectional causal relations were not revealed in the current work.

## 5. Conclusion

This preliminary study used a phenome-wide MR study of several thousand exposure variables to identify putative risk and protective factors for myopia. Our results advanced the understanding of myopia pathogenesis and may provide a potential novel prevention and treatment targets for the disease.

## Data Availability

All data produced in the present study are available upon reasonable request to the authors

https://gwas.mrcieu.ac.uk/

## Author Contributions

Conception, supervision and administration: Zenan Lin and Qi Zhang; Data curation: Junhong Jiang and Di Hu; Investigation: Junhong Jiang, and Di Hu; Methodology: Zenan Lin and Qi Zhang; Writing – original draft: Junhong Jiang and Di Hu; Writing – review & editing: Zenan Lin and Qi Zhang.

## Funding

No fund was obtained for this work.

## Conflict of Interest

The authors declare no conflict of interests.

## Ethics Approval

The used GWAS data were publicly available and approved by their original institutions. An ethics approval for the current work is not required.

## Availability of data and material

The used GWAS data were publicly available and their links were described appropriately in the paper. The codes and detailed information required to replicate the results in this work are available from the corresponding authors upon reasonable request.

## Patient and public involvement

Patients and/or the public were not involved in the design, or conduct, or reporting, or dissemination plans of this research.

## Consent for publication

Not applicable.

## Acknowledgement

The authors appreciate the efforts of the participants and investigators who made the GWAS datasets publicly available.

## Notes

### Competing Interest Statement

The authors have declared no competing interest.

### Funding Statement

This study did not receive any funding

### Author Declarations

The study used (or will use) ONLY openly available human data that were originally located at:https://gwas.mrcieu.ac.uk/datasets/

### Summary of Updates

author (Junhong Jiang and Zenan Lin) affiliations updated

