## Supplementary material for "Exploring the potential risk factors of myopia: A phenome-wide Mendelian randomization association study": Figure S1

### Lifestyle and Environment

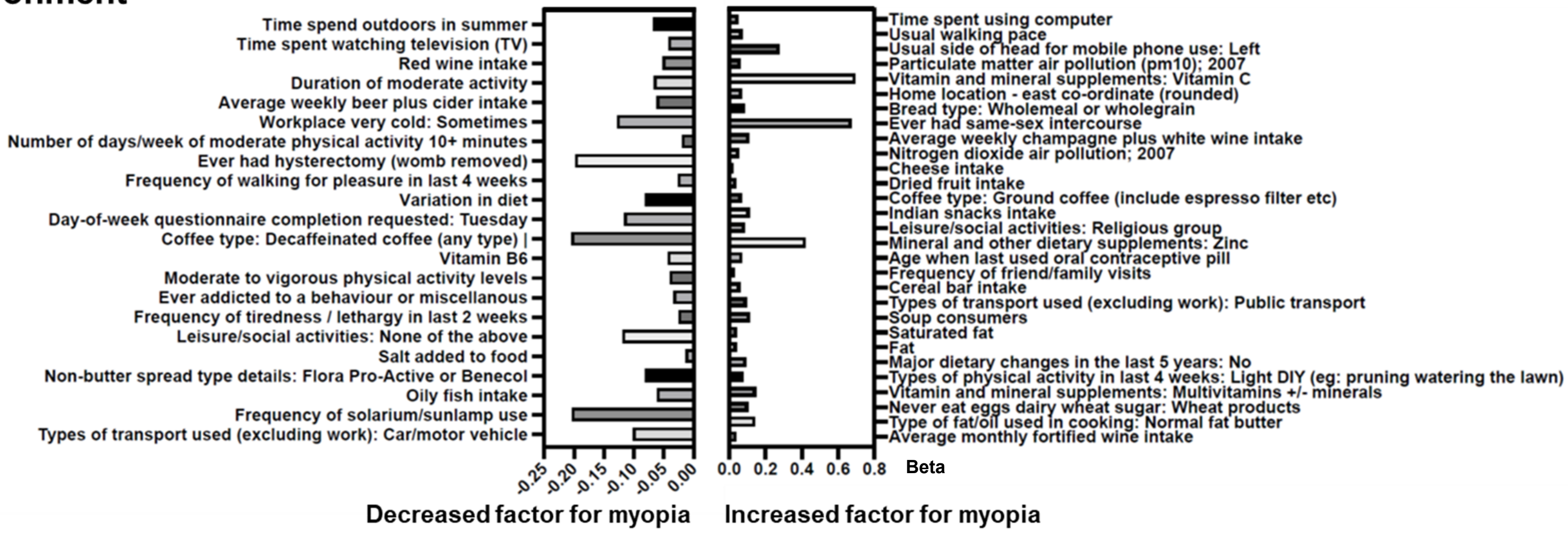


### Physical Measures

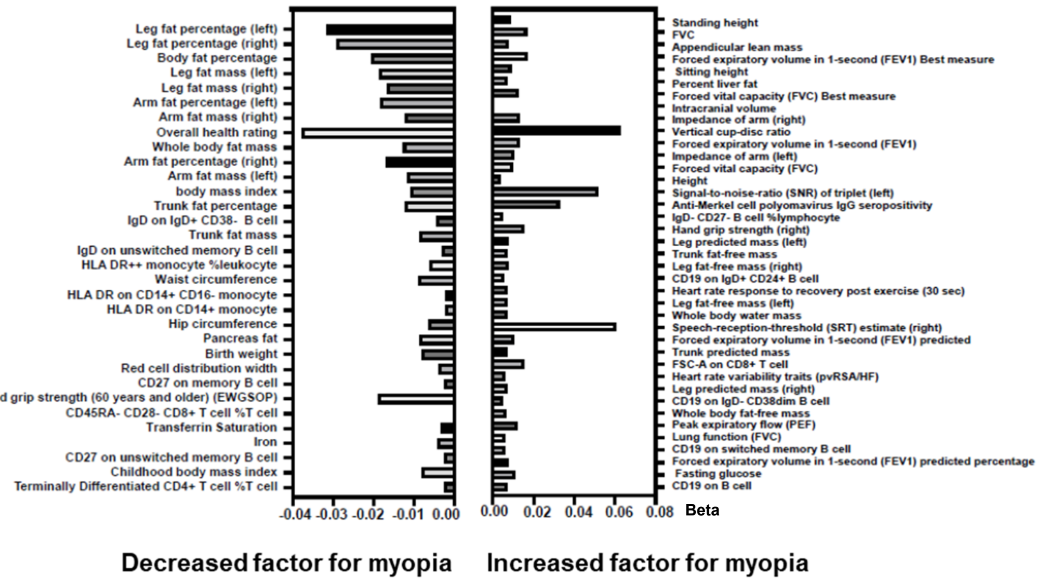


### Treatment/medications

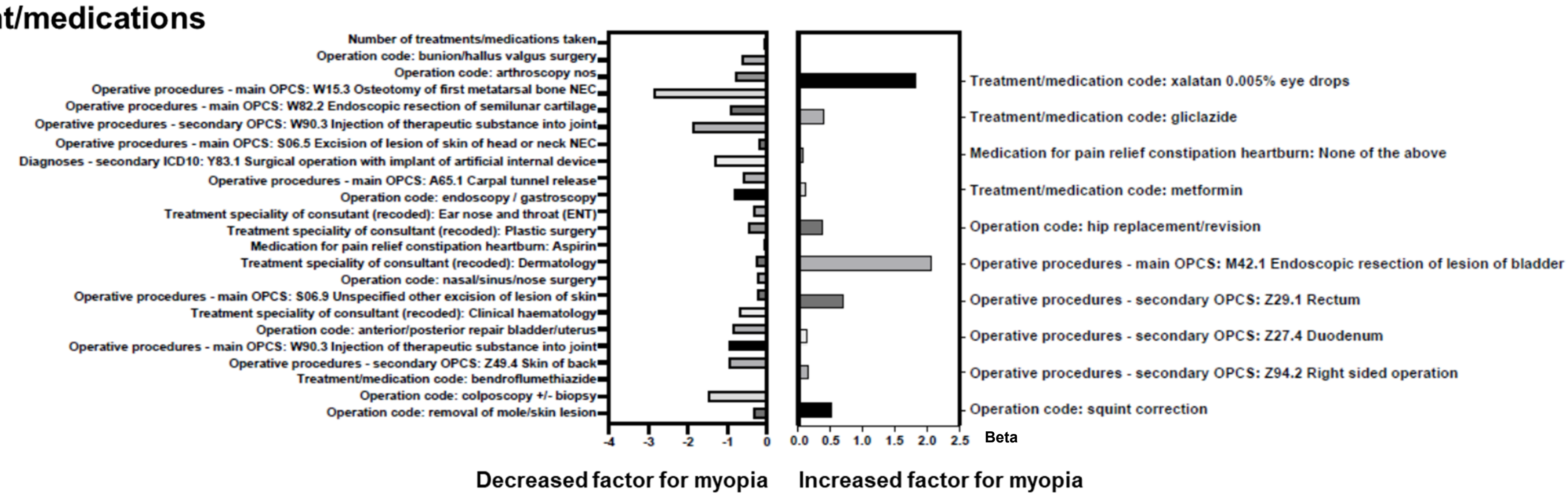


### Sociodemographics Traits

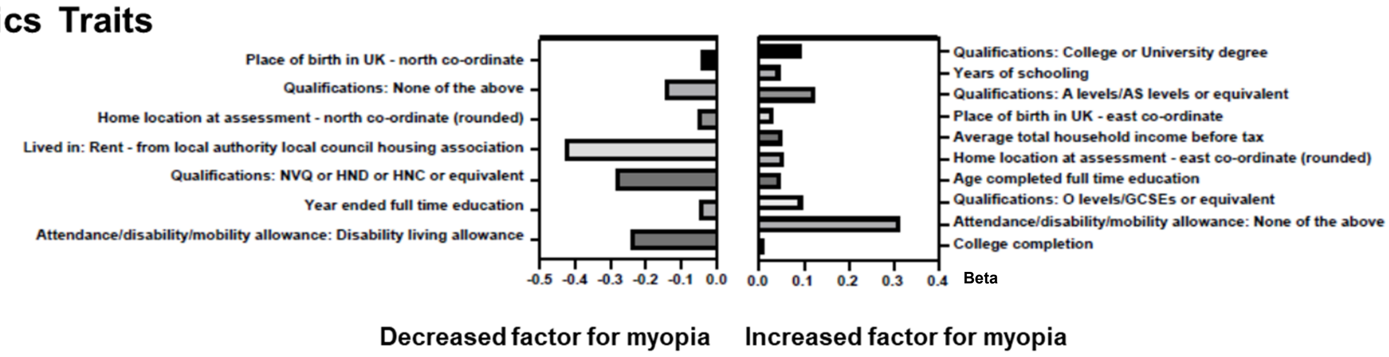


### Alcohol drinking-associated Traits

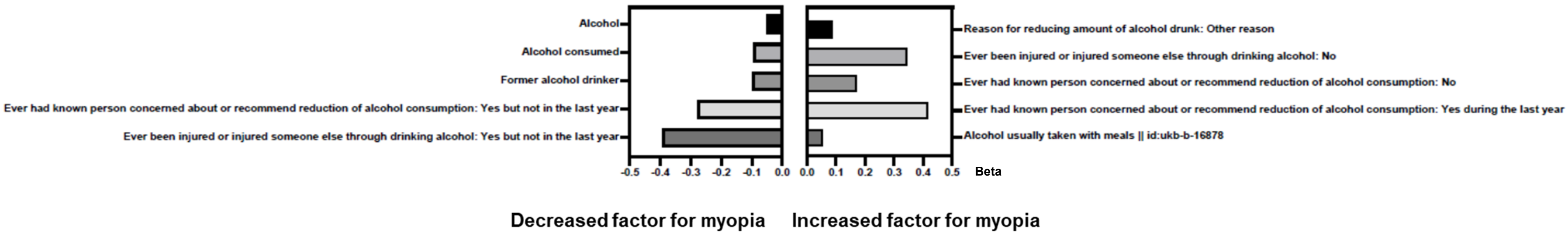


### Mental health

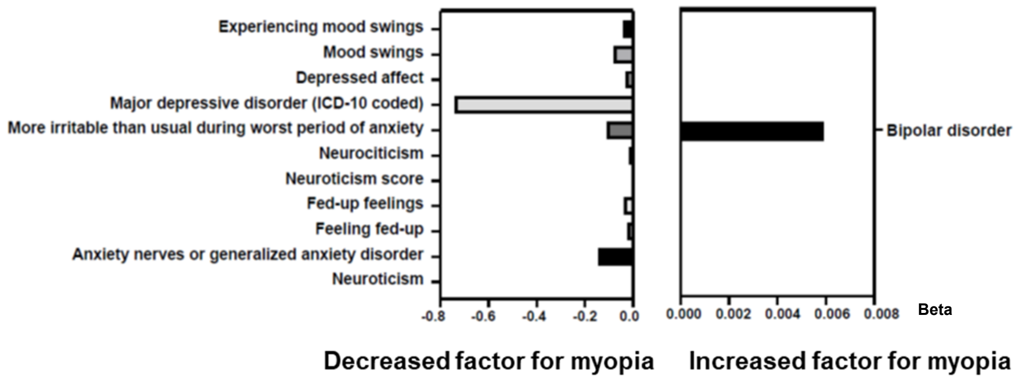
